# Antibody-mediated Immunogenicity against SARS-CoV-2 following priming, boosting and hybrid immunity: insights from 11 months of follow-up of a healthcare worker cohort in Israel, December 2020-October 2021

**DOI:** 10.1101/2021.12.15.21267793

**Authors:** Michael Edelstein, Karine Wiegler Beiruti, Hila Ben-Amram, Naor Bar-Zeev, Christian Sussan, Hani Asulin, David Strauss, Younes Bathish, Salman Zarka, Kamal Abu Jabal

## Abstract

**Background:** We determined circulating anti-S SARS-CoV-2 IgG antibody titres in a vaccinated healthcare workers (HCWs) cohort from Northern Israel in the 11 months following primary vaccination according to age, ethnicity, boosting timing and previous infection status.

**Methods:** All consenting HCWs were invited to have their circulating IgG levels measured before vaccination and at 6 subsequent timepoints. All HCWs with suspected COVID-19 were PCR tested. We described trends in circulating IgG geometric mean concentration by age, ethnicity, timing of boosting and previous infection status and compared strata using Kruskall-Wallis tests.

**Results:** Among 985 vaccinated HCWs. IgG titres gradually decreased in all groups over the study duration. Younger or previously infected individuals had higher initial IgG levels (p<0.001 in both cases); differences substantially decreased or disappeared at 7-9 months, before boosting. Pre-infection IgG levels in infected participants were similar to levels measured at the same timepoint in HCWs who remained uninfected (p>0.3). IgG GMC in those boosted 6-7 months after dose 2 was lower compared with those boosted 8-9 months after (1999-vs 2736, p=0.02).

**Conclusions:** Immunity waned 6 months post-priming in all age groups and in previously infected individuals, reversed by boosting. IgG titres decrease among previously infected individuals and the proportion of reinfected individuals in this group, comparable to the proportion of breakthrough infection in previously uninfected individuals suggests individuals with hybrid immunity (infection+vaccination) may also require further doses. Our study also highlights the difficulty in determining protective IgG levels and the need to clarify the optimal timing in 3 dose regimens

## Background

Ten months after SARS-CoV-2 was declared a pandemic, mass vaccination campaigns commenced with vaccines showing trial efficacy of over 90% against symptomatic illness [12,3]. Post-introduction empirical observational studies confirmed vaccine effectiveness against severe disease and death [4], and initially apparent effectiveness against infection [4] raised hopes of control and perhaps elimination. However, bottlenecks in production, supply and delivery and challenges in regulatory capacity meant many low- and middle-income countries remain at very low vaccination coverage [5], and vaccine hesitancy led to gaps in coverage even in countries with ready access to vaccine doses. In addition, viral variants emerged with relative immune evasion (e.g. Beta) or increased transmissibility (e.g. Delta) [6,7] that together with waning of humoral immunity from about 4 months after dose 2 [8,9] left 2-dose recipients sub-optimally protected.

In Israel, mass vaccination started in December 2020 using two doses of BNT162b2 mRNA vaccine scheduled 21 days apart as per manufacturer recommendation. As of November 2021 two-dose population coverage was 80% among persons aged 30 years and over and 75% for those aged between 16-29 [10]. In June 2021 COVID-19 community transmission ceased briefly, following which importation of the Delta variant caused the largest epidemic yet experienced in the country. Israel rapidly initiated booster doses. Experimental and observational data comparing 3 vs 2 doses, demonstrated the effectiveness of boosters against symptomatic infection with the Delta variant [11,12]. However, given the low rates of severe disease outcomes among 2 dose recipients, the absolute risk reduction is more modest, and inversely the number needed to vaccinate to avert one severe outcome is high. Thus, the appetite to introduce boosters has been variable, and as of November 2021 no other country offered universal boosting. In September 2021 the World Health Organization called for a moratorium on boosting until the end of 2021[13]. In the UK, in September 2021, the Joint Committee for Vaccination and Immunization, the independent body advising the government on vaccine policy, recommended boosting to vulnerable individuals only [14]. The duration of clinical protection conferred by the booster remains unknown, nor do we yet have a clear-cut humoral correlate of protection.

Ziv Medical Center (ZMC) is a 300-bed government regional referral hospital located in Safed, Northern Israel. Like all hospitals in the country it offered vaccination to its healthcare workers (HCW), achieving over 90% coverage. We conducted prospective serosurveillance of HCWs to evaluate trends over time in SARS-CoV-2 humoral immunity by age, vaccination, infection status and time elapsed between priming and boosting, and other predictors. Using the same cohort, we have previously published findings of vigorous anamnestic responses among previously infected single-dose recipients, and the need for second dose among individuals experiencing breakthrough primary SARS-CoV-2 infection shortly after their first dose. [15,16]

Here we describe trends in antibody-mediated immunity over 11 months following vaccination by age, ethnicity, infection status and time elapsed between priming and boosting, and compare anamnestic responses resulting from 3^rd^ dose receipt to those resulting from breakthrough infection.

## Methods

All ZMC employees were invited to participate. We verified prior infection status among consenting participants by measuring the presence of anti-Nucleocapsid (N) IgG antibodies using a highly sensitive and specific SARS-CoV-2 IgG qualitative assay (Abbott, Abbot Park, US) [17]. Workers with detectable anti-N IgG antibodies and/or documented past positive SARS-CoV-2 PCR were considered previously infected. Thereafter quantitative anti-SARS-CoV-2 Spike (S) IgG levels were measured using the LIAISON Diasorin SARS-CoV-2 S1/S2 IgG assay[17] at six time points from dose 1; t_1_: 21 days (range 15-35 days), t_2_: 51 days (range 41-65 days), t_3_: 100-150 days, t_4_: 151-210 days, t_5_: 211-270 days and t_6_: 271-310 days. Where the IgG level reading reached the maximum, serial dilutions were performed in order to obtain a precise quantitative value. HCWs were asked to report any arising symptoms. Those whose symptoms were consistent with the standard clinical case definition of COVID-19 were tested by rtPCR. Individuals with a positive PCR test were classified as infected post-vaccination (breakthrough infection). Antibody levels were reported using geometric mean concentration (GMC) alongside 95% confidence intervals (95% CI). We used log-GMC when reporting trends graphically. anti-S IgG GMCs were reported by strata defined by number of vaccine doses received, infection status (never infected, infected prior to vaccination, infected after full vaccination), age (according to age at recruitment), ethnicity and timing of boosting. We tested to reject the null hypothesis of no difference in GMC across strata using Kruskall Wallis tests. In order to determine any differences in immunogenicity by age and ethnicity we restricted analysis to never infected individuals who had received at least 2 doses of vaccine. We restricted the ethnicity analysis to individuals aged 35-54 because of the higher proportion of older HCWs in the Jewish group compared with others. It is worth noting that the number of individuals providing a blood sample at each time point varied (range: 324-646) and therefore the GMC at each time point is based on a different number of individuals. The study was approved by ZMC’s ethics committee (0133–20-ZIV).

## Results

Of 1500 employees, 985 consented to take part in the study, received at least one dose of vaccine and had at least one serological test post vaccination. Of these, 86 received only a single dose, 141 received 2 doses and 758 received 3 doses (Table 1). The median time between doses 1 and 2 was 21 days, and 223 days between doses 2 and 3. HCWs who received a single priming dose (generally because of previous infection) and a second dose more than 6 months after the first were considered boosted. One hundred and eighteen HCWs were infected prior to vaccination, of which 7 (5.9%) were re-infected after vaccination. Of the 856 participants who received at least two doses and were seronegative at the initiation of vaccination, 82 (9.6%) were infected after initiating their vaccine course of which 40 (4.7%) 30 days or more after receipt of dose 2. There was no association between age group and the incidence of breakthrough infection (p=0.26). Participants of all ages, genders and ethnicities represented in the general population of Israel were represented in the sample (Table 1).

**Table 1.** characteristics of participants.

| Table 1. characteristics of participants |  |  |
| --- | --- | --- |
|  | n | % |
| **Number of priming doses** |  |  |
| one priming dose | 86 | 9 |
| two priming doses | 899 | 93 |
| **Booster** |  |  |
| Yes | 758 | 77 |
| No | 227 | 23 |
| **Infection status** |  |  |
| previously infected | 118 | 12 |
| infected post vaccination | 93 | 9 |
| never infected | 779 | 79 |
| **Age** |  |  |
| Under 35 years old | 258 | 26 |
| 35-44 years old | 238 | 24 |
| 45-54 years old | 224 | 23 |
| 55+ | 243 | 25 |
| unknown | 22 | 2 |
| **Ethnicity** |  |  |
| Jewish | 437 | 44 |
| Christian | 77 | 8 |
| Muslim | 110 | 11 |
| Druze | 76 | 8 |
| Circassian | 6 | 1 |
| unknown | 279 | 28 |
| **Gender** |  |  |
| female | 613 | 62 |
| male | 372 | 38 |

We observed a decrease in circulating IgG levels over the follow up period overall and in all predefined subgroups. At t_1_, compared with never infected individuals, those previously infected (referred to in the litterature as having hybrid immunity or “superimmunity” [18]) had 13-fold higher GMC (876.6 vs 63.9, p<0.0001). Among the same individuals the fold-difference at t_4_ (5-7 months post dose 1) was 1.9 (268.4 vs 139.1, p<0.001). At t_5_ there was no statistically significant difference in GMC between the two groups although the number of previously infected individuals with available data at this time point was very small (n=4). Among never-infected participants, younger age was associated with higher GMC post dose 1(t_1_) (table 2, p<0.001) but the difference in GMC was barely significant by t_5_ (7-9 months post dose 1 but prior to dose 3, table 2, p=0.05). There was no association between GMC and ethnicity among never infected, fully vaccinated individuals at any time point.

**Table 2.**
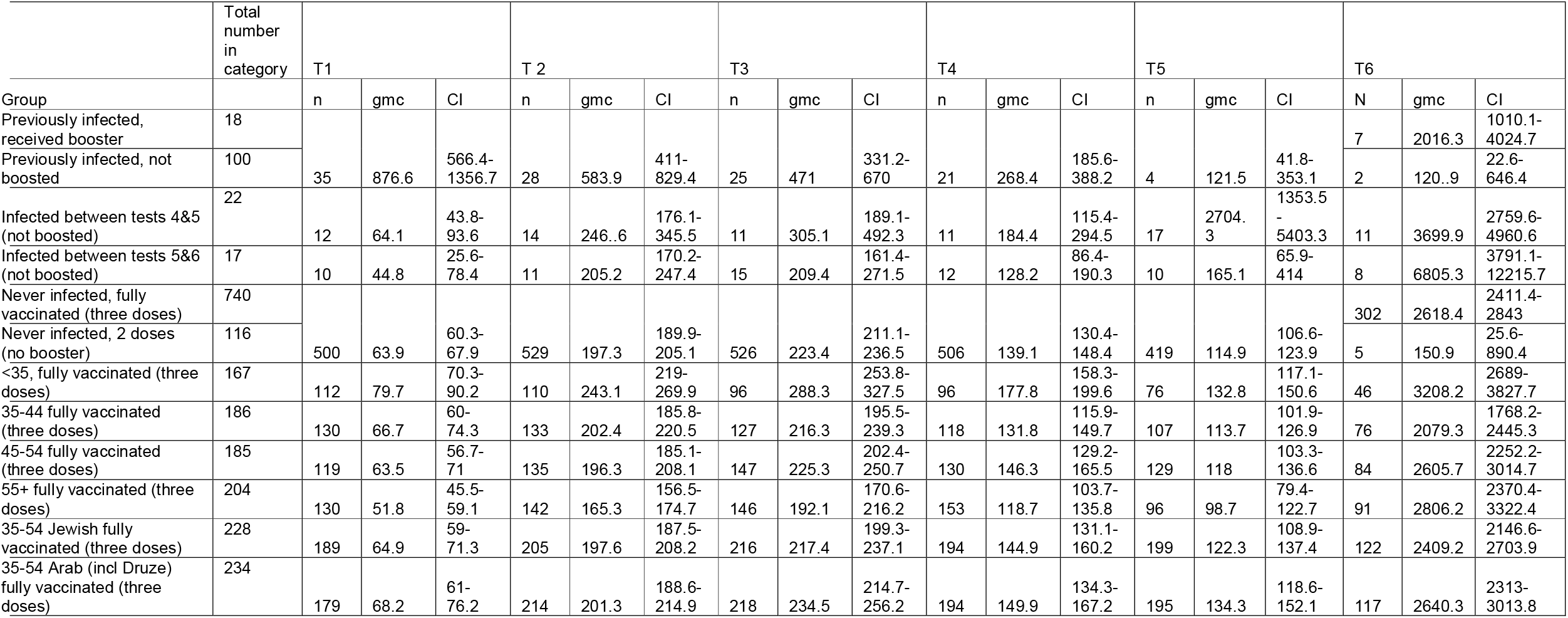
**Geometric mean concentration of anti-SARS-CoV-2 spike IgG antibodies among healthcare workers in the 11 months following vaccination against COVID-19, Israel, December 2020 to October 2021**

Of the 899 HCW who received ≥2 doses, 40 (4.4%) were confirmed positive on PCR between 30 days after dose 2 and before dose 3. Of those, 4 tested PCR positive prior to t4, 20 had a positive PCR test between t_4_ and t_5_ and 16 between t_5_ and t_6_. Among those infected after vaccination, IgG GMC just prior to infection was not different than among those who remained uninfected at the same time point (184 vs 139, p=0.3 for those infected between tests 4 and 5, 165 vs114, p=0.9 for those infected between tests 5 and 6). The 40 individuals experiencing breakthrough infections were younger than never infected HCWs (mean age 39 vs 45 years old, p<0.002).

Of the 302 never infected HCWs who received dose 3 and were tested 1-2 months afterwards, t_6_ GMC (1-2 months post dose 3) was 2618 (95%CI 2411-2843), while among the 21 non-boosted individuals infected after dose 2 for whom data was available, GMC was significantly higher (4213, p< 0.001, Fig 2). Among those never-infected, all age groups saw an increase in IgG levels 18-fold or more post boosting. GMC in those boosted 6-7 months after dose 2 was lower compared with those boosted 8-9 months after dose 2 (1999-vs 2736, p=0.02).

**Figure 1.**
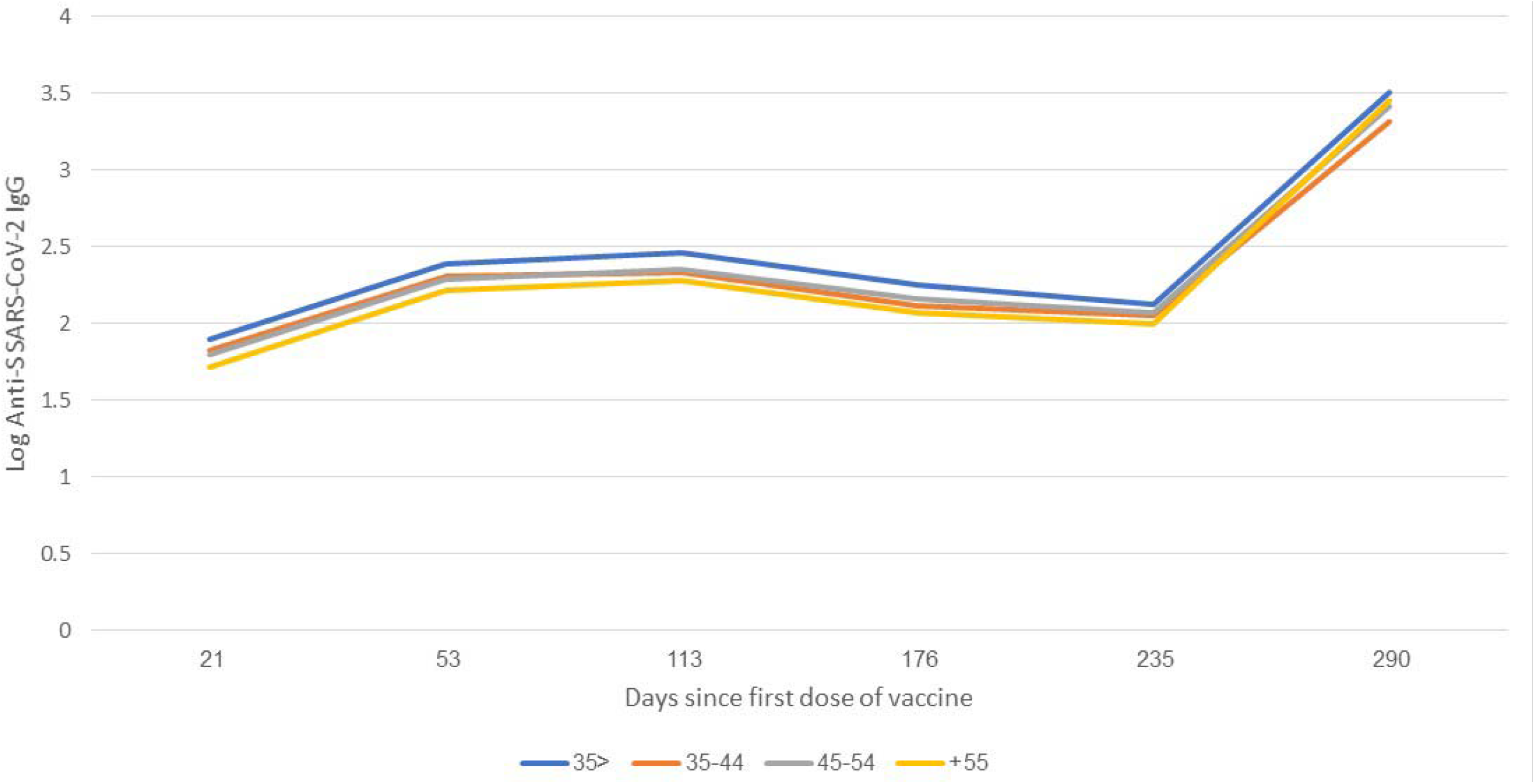
Anti SARS-CoV-2 S IgG Geometric mean concentration (log) according to age, Israel, January-October 2021

**Figure 2.**
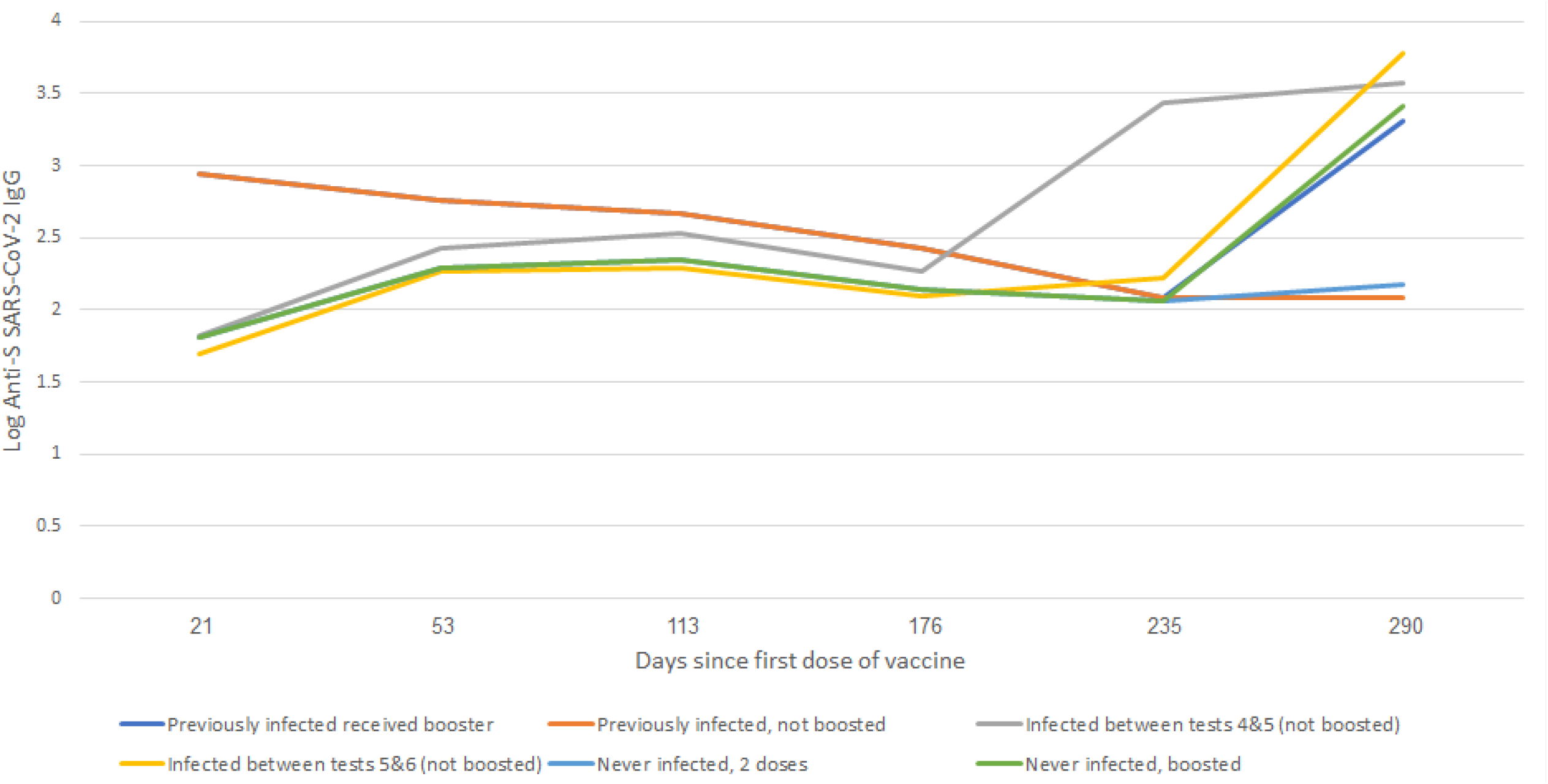
Anti SARS-CoV-2 S IgG Geometric mean concentration (log) according to infection status, Israel, January-October 2021

## Discussion

Our convenience cohort provided a well representative setting in which to monitor serologic responses over time. Consistent with other observational data[8], we found that IgG titres begin waning from about 6 months post dose 2 in all age groups, and that this phenomenon occurs irrespective of previous infection status. We also found that despite initially higher GMCs in younger individuals, after 6-7 months differences were much smaller or no longer apparent, suggesting that all age groups might require boosting to achieve optimal protection. Previously infected individuals, who had IgG levels one order of magnitude higher than those never infected after one dose[15] also saw their circulating IgG levels drop at 6-7 months, with levels less than twice as high as those never infected. The proportion of reinfections among individuals infected pre-vaccination was comparable to the proportion of breakthrough infections among never infected individuals who received two doses. These findings suggest that, in line with other observational studies[19], hybrid or super-immunity (natural immunity boosted by vaccination), wanes and may eventually need boosting as well, at least if decision making is based on circulating IgG levels. Observational data has shown a high effectiveness of boosting against infection and severe disease[11,12] including against the recently emerged Omicron variant[20]. The IgG levels achieved after boosting were one order of magnitude higher than after the priming course and close to levels achieved in those infected after the priming course. Our data do not allow to estimate duration of protection and no robust real world or modelling studies that estimate duration of protection are available yet. The large fold-increase in circulating IgG following infection among vaccinated individuals may also have diagnostic value where it is not possible or practical to swab individuals for PCR tests during the narrow window of opportunity that the PCR modality offers.

We have demonstrated previously no difference in GMC by ethnicity following a single dose of vaccine. In the present study we found that this remains consistent after subsequent doses. This findings matter because risk of infection and disease was indeed associated with ethnicity in Israel and elsewhere, both before and after national introduction of COVID-19 vaccine [21,22,23].

Our study also highlights the limits of using circulating IgG to determine immunogenicity. Anti-S IgG GMCs measured just prior to infection among individuals who became infected after dose 2 were not significantly different than uninfected individuals at the same time point. Infected individuals had high circulating IgG levels just prior to infection (>100 AU/ml on average, much higher in some individuals) and would have been considered strongly positive on any routine serology test. These elements suggest circulating IgG levels are not a robust predictor of protection against infection or disease and it is not currently possible to easily determine correlates of protection for COVID-19. Evidence demonstrates the persistence and importance of cellular immunity, both B and T cell[24,26,25]. Confirming protection following vaccination or infection cannot solely rely on circulating IgG titres and requires other measures of immunity such as functional assays, or B cell and T cell assays, none of which are routinely available for diagnostic purposes. Our study also suggests that the timeframe in which the booster is offered in Israel-6 months after the second dose-triggers a large anamnestic response. However later boosting was associated with higher IgG levels. It is unclear at this stage to what extent these differences are clinically relevant in terms of effectiveness or how they will impact on the duration of protection. A better understanding of how IgG levels correlate with protection followed by head to head studies of different boosting schedules to optimise protection longevity are required, especially where new variants continue to emerge and calls for further doses beyond a single booster are beginning to be made.

The decrease in IgG levels in the cohort described in this study occurred during a time of increase in the incidence of reported COVID-19 infection in Israel[8] but also at a time of a shift in the dominant circulating strain in Israel from Alpha to Delta. It is therefore a challenge to distinguish the effects of declining immunity from those of higher infectivity attributable to novel variants. In addition, while waning immunity has caused vaccine effectiveness against infection to decrease from over 90% to 50-60%27], the decrease in effectiveness against severe outcomes such hospitalization and death is much less pronounced [28]. While our study supports widespread boosting in all age groups from the immunogenicity perspective, the public health benefit of boosting should be balanced against priming previously unvaccinated individuals, both at the national and global levels, when formulating boosting policies.

Repeated blood sampling in the cohort was challenging. The number of latter tests was small, particularly within strata. We caution against drawing inference from later subgroup comparisons. Secondly, PCR testing only occurred upon report of symptoms which likely underascertained true infection incidence with potential misclassification of infected asymtpomatic participants as never infected. Though we did not observe increases in titres unexplained by either vaccination or reported symptoms. Finally though we compared titres, we did not measure neutralizing ability.

Our study demonstrates antibody waning and high post-boosting IgG levels in all age groups, suggesting widespread boosting policies may be beneficial, although this needs to be substantiated by effectiveness studies going forward. The need for such policy becomes more urgent with the emergence of strains such as Omicron that likely requires much higher antibody titres for neutralisation in order to achieve protection29,30]. Our data suggest that immunological waning occurs in vaccinated, naturally infected, and infected-then-vaccinated groups, regardless of age and ethnicity. Ongoing detailed large observational cohorts that measure antibody function and have sufficient clinical outcome incidence will help clarify to what extent, after how long and in terms of which variants, these individuals are again at risk. We continue to monitor anti-S titres in order to determine the durability of boosted immune responses by age, infection history and interval between priming and boosting.

## Data Availability

All data produced in the present study are available upon reasonable request to the authors

